# An analysis of school absences in England during the Covid-19 pandemic

**DOI:** 10.1101/2021.02.10.21251484

**Authors:** Emma Southall, Alex Holmes, Edward M. Hill, Benjamin D. Atkins, Trystan Leng, Robin N. Thompson, Louise Dyson, Matt J. Keeling, Michael J. Tildesley

## Abstract

The introduction of SARS-CoV-2, the virus that causes COVID-19 infection, in the UK in early 2020, resulted in the UK government introducing several control policies in order to reduce the spread of disease. As part of these restrictions, schools were closed to all pupils in March (except for vulnerable and key worker children), before re-opening to certain year groups in June. Finally all school children returned to the classroom in September. In this paper, we analyse the data on school absences from September 2020 to December 2020 as a result of COVID-19 infection and how that varied through time as other measures in the community were introduced. We utilise data from the Educational Settings database compiled by the Department for Education and examine how pupil and teacher absences change in both primary and secondary schools.

Our results show that absences as a result of COVID-19 infection rose steadily following the re-opening of schools in September. Cases in teachers were seen to decline during the November lockdown, particularly in those regions that had previously been in tier 3, the highest level of control at the time. Cases in secondary school pupils increased for the first two weeks of the November lockdown, before decreasing. Since the introduction of the tier system, the number of absences owing to confirmed infection in primary schools was observed to be significantly lower than in secondary schools across all regions and tiers.

In December, we observed a large rise in the number of absences per school in secondary school settings in the South East and Greater London, but such rises were not observed in other regions or in primary school settings. We conjecture that the increased transmissibility of the new variant in these regions may have contributed to this rise in cases in secondary schools. Finally, we observe a positive correlation between cases in the community and cases in schools in most regions, with weak evidence suggesting that cases in schools lag behind cases in the surrounding community. We conclude that there is not significant evidence to suggest that schools are playing a significant role in driving spread in the community and that careful monitoring may be required as schools re-open to determine the effect associated with open schools upon community incidence.

## 1 Introduction

In late 2019, a novel strain of coronavirus, now known as SARS-CoV-2, emerged in Wuhan, China [1, 2]. Over the next few months, this virus spread around the world, with the World Health Organisation declaring a global pandemic on 11th March 2020 [3]. Upon infection with the virus, individuals can develop COVID-19 disease. In some instances, infected people do not develop symptoms or are only mildly infected, with symptoms including a dry cough, a fever, shortness of breath and a loss of taste and smell [4–6]. However, in more serious cases, predominantly in the elderly and those with underlying health conditions, hospitalisation and admission to intensive care may be required, with many of these individuals dying as a result of their infection [7–10].

In the UK, the government began to introduce control policies in March 2020 in order to prevent the spread of infection. These included the closing of all pubs, restaurants and non-essential shops on 20th March, as well as the closing of schools [11] to all pupils except for vulnerable children or those with key worker parents. On 23rd March, the UK entered full lockdown, whereby people were only allowed out of their house for essential shopping, medical treatment, essential work and one form of exercise per day [12].

As cases in the UK started to decline in May, there was a push for a staggered return to the classroom and on 1st June, primary schools re-opened for pupils in reception, year 1 and year 6 (typically ages 4-5, 5-6 and 10-11), whilst from 15th June, secondary schools began to re-open for pupils in years 10 and 12 only (typically aged 14-15 and 16-17; [13]). All other children were required to learn from home for the remainder of the academic year.

All children returned to school in England from 1st September, but with non-pharmaceutical interventions (NPIs) in place [14]. In many instances this involved staggered drop off and pick up times for children, mandatory wearing of masks in playgrounds for parents and in indoor settings excluding classrooms for secondary school children, as well as advice to parents not to congregate outside the school gates. Additionally, children and staff were placed into “bubbles” in order to minimise the risk of large scale spread. In primary schools, these bubbles were introduced either at the level of the class or the year group, whilst in most secondary schools the entire year group would typically form a bubble, owing to significant mixing across different academic subjects [15]. If a pupil or staff member within a bubble tested positive for COVID-19, all others within the bubble were required to isolate for 14 days and were advised to seek a test if they started to develop symptoms.

During October 2020, as cases began to rise, the government introduced a regional three tiered system in order to control disease spread [16, 17]. Each region of the country would be placed into a tier dependent upon the local incidence, the effective reproduction number and the local hospital occupancy. In all tiers, the “rule of six” was in place, which prohibited mixing in groups of more than six people, but as regions escalated through the tiers, the settings in which the individuals could meet outside their household became more restricted in an attempt to curb the spread of infection. When the tiers were introduced on 14th October 2020, the majority of the South of England and the Midlands were in tier 1, the lowest level of control, whilst many parts of the North were first placed into tier 2, with some regions escalating further into tier 3 a few days later. By the end of October 2020, cases of COVID-19 were rising across the country in a concerning way [18, 19] and the government announced that a four week lockdown would be introduced in England from 5th November 2020. However, schools remained open during this lockdown, as it was decided that the need for children to remain in education outweighed the risks associated with schools remaining open.

The emergence of a new, more transmissible variant, B.1.1.7, in the South East of England during this period contributed to a significant increase in spread towards the end of the year, particularly in the South East, Greater London and the East of England [20]. It became apparent that the November lockdown in England had not been sufficient to bring the reproduction number (R) below 1 and that more action would be needed urgently to avoid the National Health Service becoming overwhelmed in the new year. Over the Christmas period, there was significant debate over the need for another national lockdown and whether or not this lockdown should include the closure of schools. Finally, on 4th January, the government announced that a new lockdown would be introduced in England with immediate effect and that schools would again be closed to all children except vulnerable children and those with key worker parents [21].

The closing of schools was brought in as a necessary measure on 20th March 2020 and again on 4th January 2021, owing to the need for a significant tightening of restrictions in order to bring the R number below 1. However, decisions to close schools need to balance the risk associated with transmission of SARS-CoV-2 with the negative impact of school closures upon children’s educational needs and their health and well being. The evidence to date strongly indicates that the vast majority of children are only mildly affected by the disease and the mortality rates are extremely low [22, 23]. There has been a higher level of uncertainty regarding children’s role in transmission of the virus [24, 25] and the effect that the closing of schools will have upon the reproduction number, though when schools are open there is the potential for increased mixing of parents which may contribute to transmission. In order to ensure that children can return to the classroom as quickly but safely as possible, it is important to understand the risk in both primary and secondary settings and how that risk may vary depending on the incidence of COVID-19 in the local community.

In this paper, we analyse data from the Department for Education on school absences in primary and secondary settings throughout the pandemic, and how the level of absences were dependent upon the current state of the pandemic and the level of controls that were in place in the wider community at the time. Owing to the restrictions that were in place before the summer vacation and the fact that schools were only open to key worker children, vulnerable children and specific year groups at that time, the overall attendance at school showed significant variability during 2020 (Fig. 1). We focus here upon the autumn term, from September to December 2020, when all children were in school, in order to provide evidence regarding the risk associated with schools remaining open during this time.

**Fig. 1:**
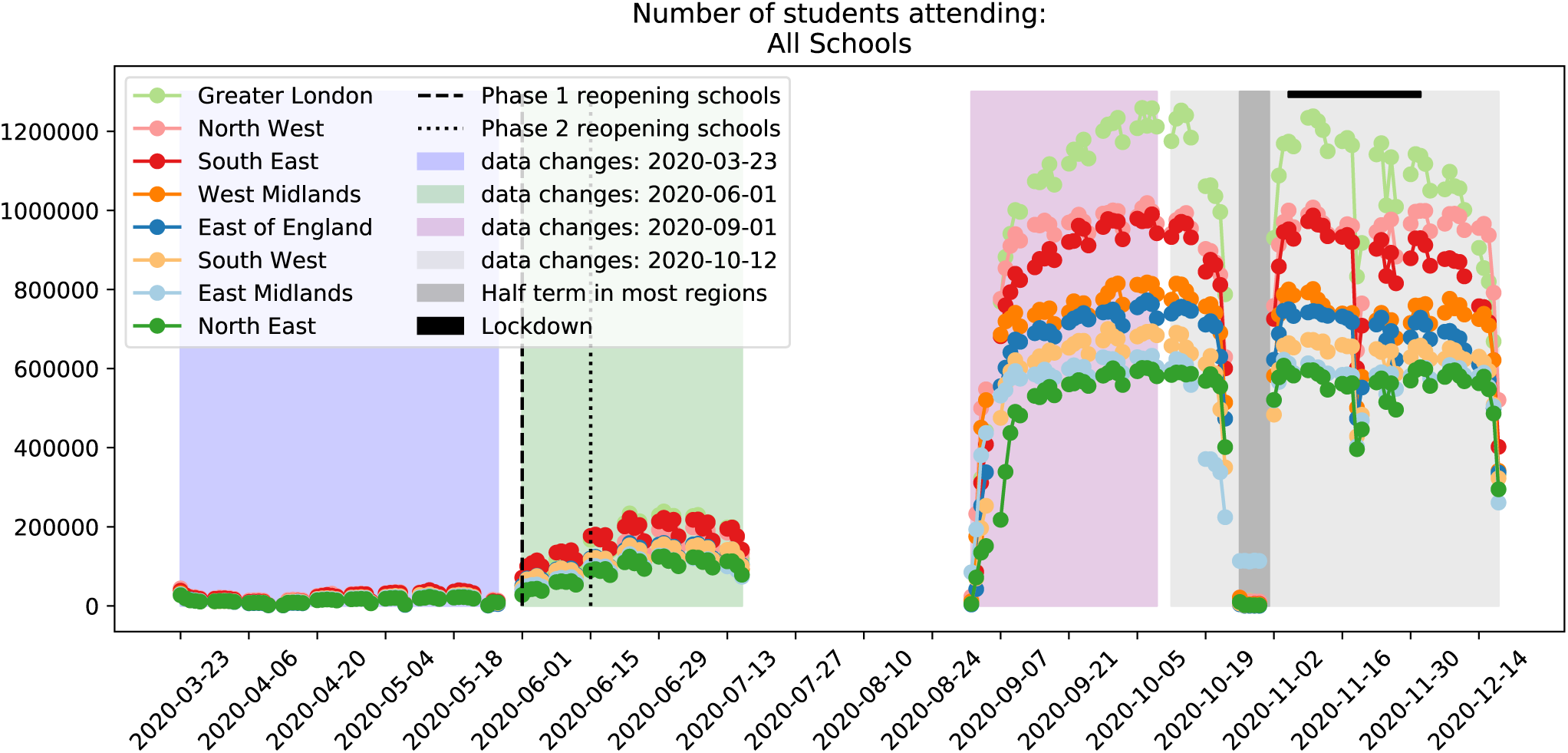
Number of students attending schools between 22nd March 2020 and 17th December 2020.

## 2 Methods

### 2.1 Data Sources

The Department for Education: Educational Setting Status data [26] were extracted over four time periods: 22nd March 2020 to 30th May 2020; 1st June 2020 to 16th July 2020, 1st September 2020 to 10th October 2020 and 11th October 2020 to 17th December 2020. A database for each extraction contains the status of each school on each day throughout the extracted period, including quantitative and qualitative records for attendance and absences. Available in all records were the school ID number, time stamp and the number of pupils in attendance. Details summarising the changes made to the databases throughout the time period we analysed are given in Table 1. In this paper, we primarily consider the data collected in the Autumn term (1st September 2020 to 17th December 2020) when all pupils on roll were eligible to attend school. It is important to note that during the first half of this time frame (1st September 2020 to 10th October 2020), records of teacher absences were limited, with less than 1% of schools submitting this information.

**Table 1:**
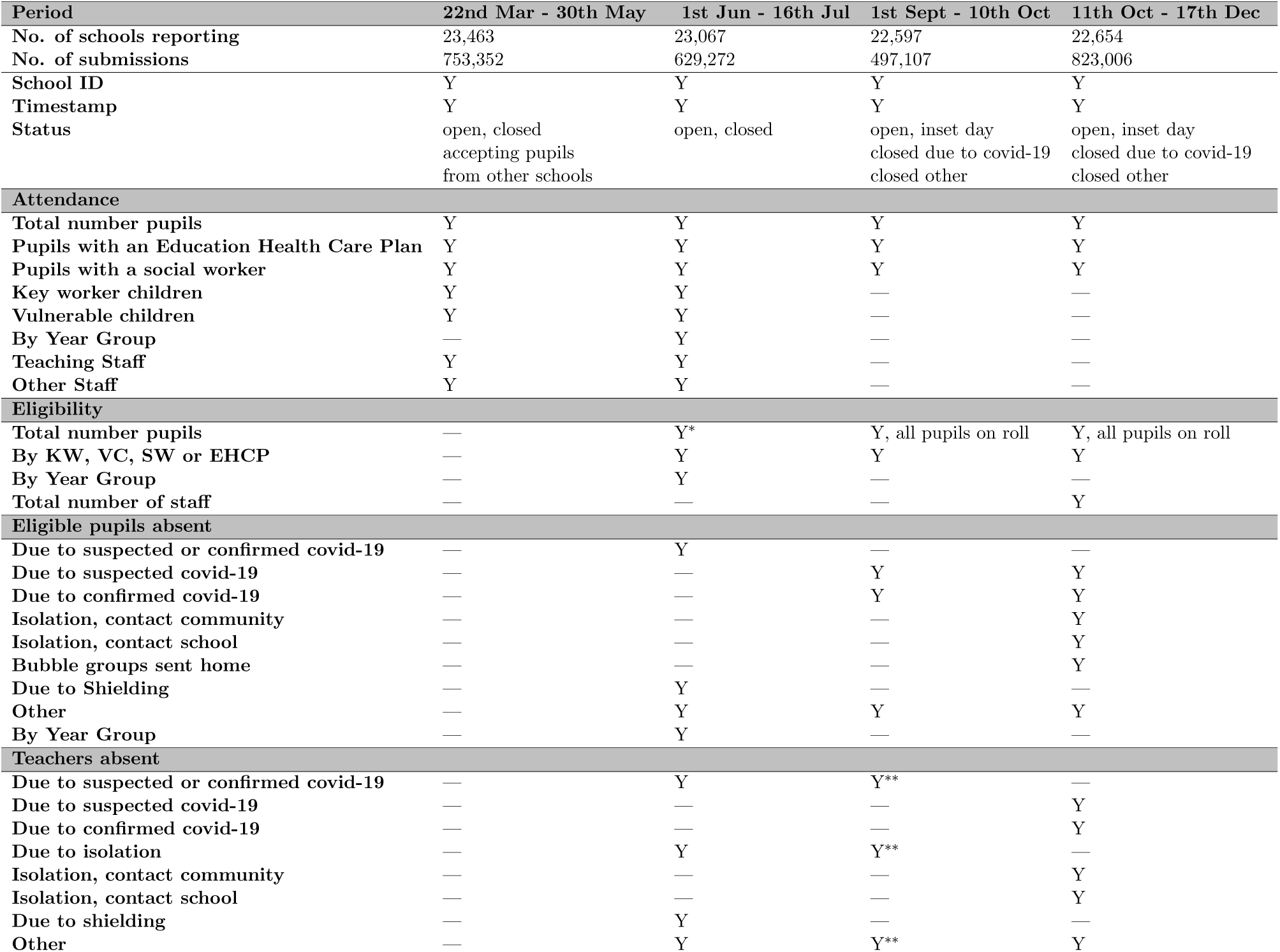
Data elements available in Department for Education: Educational Setting Status data. Notes: * - after 22nd June 2020 entry is unreported; ** - only 203 schools (out of 22,597) reported teachers absences; “Y” indicates that data are available whilst “—” indicates that data are not available

From the school URN number (ID), the location of each school including postcode, Local Tier Local Authority (LTLA) and regional information was extracted from the government database [27]. Additionally, phase of education (e.g. primary or secondary) and establishment type (e.g. state, private, academy) were accessible for each school.

### 2.2 Data Processing

We cleaned the data by removing any rows with missing date values and smoothing the pupil roll and teacher roll over the term 1 period. We approximated the total number of pupils on roll at each school by taking the maximum of the daily number of pupils on roll at each school over term 1. This was performed to smooth out any small changes over the term, particularly to remove the drop in eligibility during half-term and the usual staggered opening of early years schooling in September, when reception children typically first go to school in a phased manner over the first two weeks of term. Teacher roll was not available until 11th October 2020 (as described in Table 1). We approximated the teacher-roll prior to 11th October 2020 by taking the maximum recorded in each school between the 11th October 2020 to 17th December 2020 period, assuming that the teacher roll is constant over a school term.

### 2.3 Data Analysis Methods

The data were aggregated spatially by summing over each LTLA or region. It is these aggregated values that we used for our spatial analyses, rather than considering each school individually within a region. Additionally, we also grouped the data by the current control policies that were in place in the corresponding LTLA – this was computed by aggregating over all schools under each tier allocation over time, whereby each school was categorised by the tier of its LTLA each day. To study the spatial-temporal patterns, we compute and map community case percentages as the proportion of positive tests from the Pillar 2 PCR test data for each LTLA, averaged over the five day school week; teacher and pupils percentages were calculated based upon the proportion of teachers and pupils who are absent from school due a positive test in each LTLA, averaged over the five day school week. Relative incidence thresholds were calculated in the first week of November by assigning the top 10% of LTLAs to the ‘Very high’ category, the 75th-90th percentile as ‘High’, the 50th-75th percentile as ‘Medium’ and the remainder ‘Low’. We then used the same threshold values in all subsequent weeks, to allow for comparison across weeks.

To assess the impact of the new variant, B.1.1.7, we considered specific regions which, at the time, had been differentially impacted by the new variant. London and Kent were chosen due to the high number of new variant compatible cases reported, while Devon and the West Midlands were chosen due to having had fewer reported cases compatible with the new variant. Additionally, Devon and West Midlands had differing tier statuses on the 3rd December 2020, with Devon in tier 2 and the West Midlands in tier 3.

We applied two approaches to assess the possible impact of the B.1.1.7 variant on school absences: (i) inspect the distribution of student absences due to a confirmed covid-19 case on a day in November 2020 and a day in December 2020; (ii) analyse lagged correlations between absences and community cases.

We inspected the distribution of student absences due to a confirmed covid-19 case on 4th November 2020 and 16th December 2020 in each of the four regions. Explicitly, for each region, we identified the number of students absent in each school due to a confirmed positive test on the specified days and observed how these absences per school were distributed on each day.

To study lagged correlations between absences and community cases, we calculated the Pearson’s correlation coefficient between the number of community cases in an LTLA on one day, and the number of pupil absences due to a confirmed positive covid-19 test on another day. We considered discrete daily lags from [-10, 12], with a lag of +*k* referring to the correlation between cases in the community on day *t−k* and absences in school due to a confirmed covid-19 test on day *t*. We considered primary and secondary schools separately, with a single school on one day within the specified region corresponding to a single data point.

## 3 Results

### 3.1 Temporal variation in absences in schools by region

We analysed the total number of confirmed cases in schools in all regions (Fig. 2). Cases in pupils steadily increased in all regions following the return to school in early September. We note that, owing to a change in the data recording system in mid October, a distinct increase in the number of secondary school pupils absent is observed across all regions. We believe that this is an artefact of an alteration in the data recording system rather than a true rise in absences in that week. Following half term in late October, confirmed cases in pupils continued to rise, noticeably in secondary schools. Throughout this period, the percentage of confirmed cases in secondary school pupils was much higher than in primary schools. Cases were seen to reduce in all regions two weeks after the introduction of lockdown in November. In December, cases in secondary school students in Greater London increased markedly (Fig. 2(b), light green line), but in other regions, particularly those in tier 3 such as the West Midlands and the North West (Fig. 2(b), orange and pink lines), cases continued to decrease, indicating that a reduction of spread in the community may have resulted in a reduction of cases in schools.

**Fig. 2:**
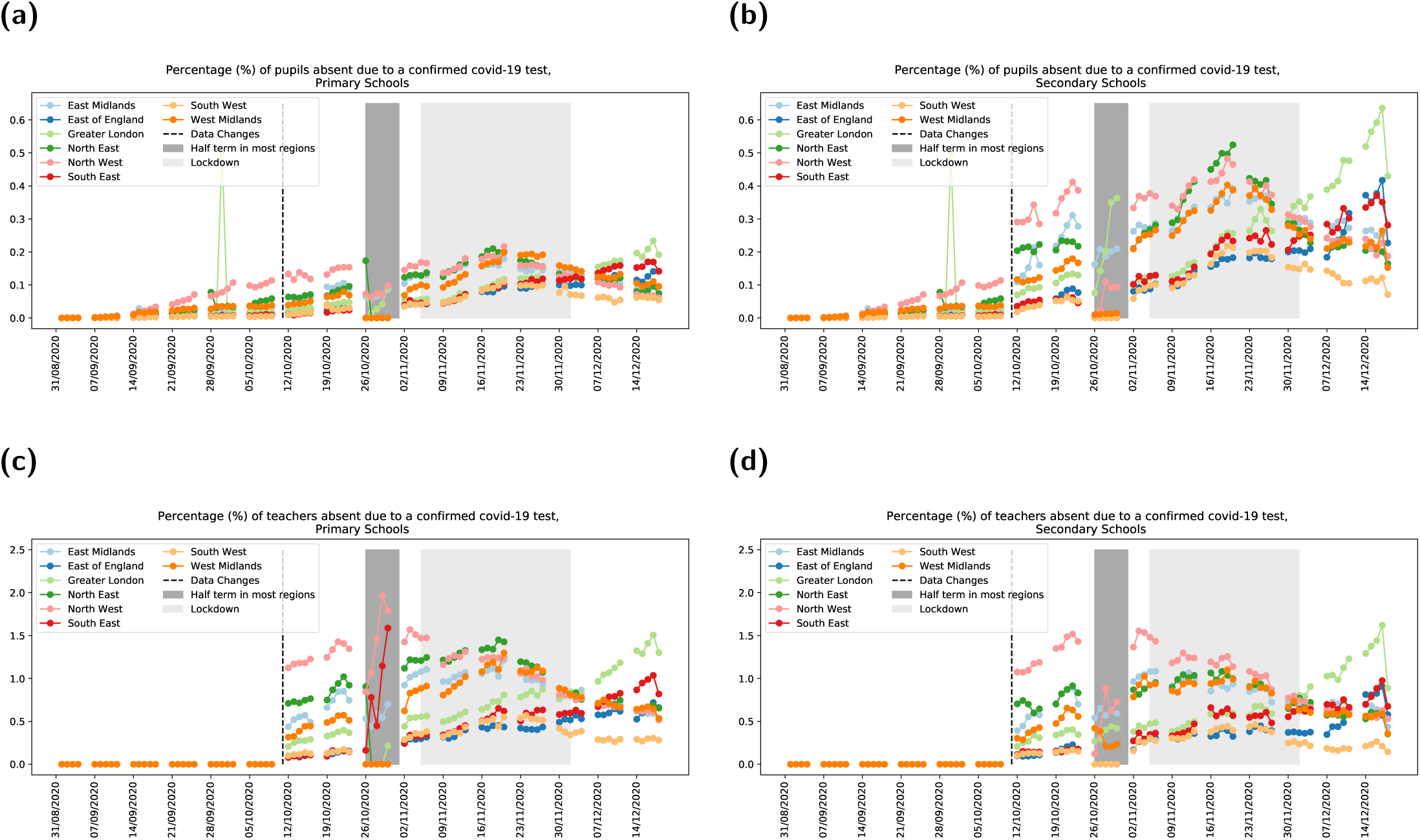
Percentage of study population recorded as a confirmed case, stratified by region. For each panel, we display the number of cases by date and by region, from 1st September 2020 to 17th December 2020. Cases in teachers were not recorded in the data prior to the 14th October 2020, when the data outputs from DfE were updated (light grey shaded region). The half term week for most of England is shown by the dark grey shaded region whilst the light grey shaded region represents the national lockdown in England which commenced on 5th November. **(a)** Pupils in primary schools. **(b)** Pupils in secondary schools. **(c)** Teachers in primary schools. **(d)** Teachers in secondary schools.

Confirmed cases in teachers declined throughout November in regions under greater restrictions prior to lockdown (North West, North East, West Midlands), compared to a slight increase in lower tier control regions. We did not observe a marked difference between the percentage of confirmed cases in teachers in primary and secondary schools (Figs. 2(c) and 2(d)). The number of cases in teachers increased in Greater London and the East of England in December, but at a lower rate than in secondary school pupils. For all regions, in both primary and secondary schools, we find a strong correlation between cases in pupils and teachers, with a larger number of cases in students in secondary schools but no evidence of increased risk to teachers in this setting (Fig. 3).

**Fig. 3:**
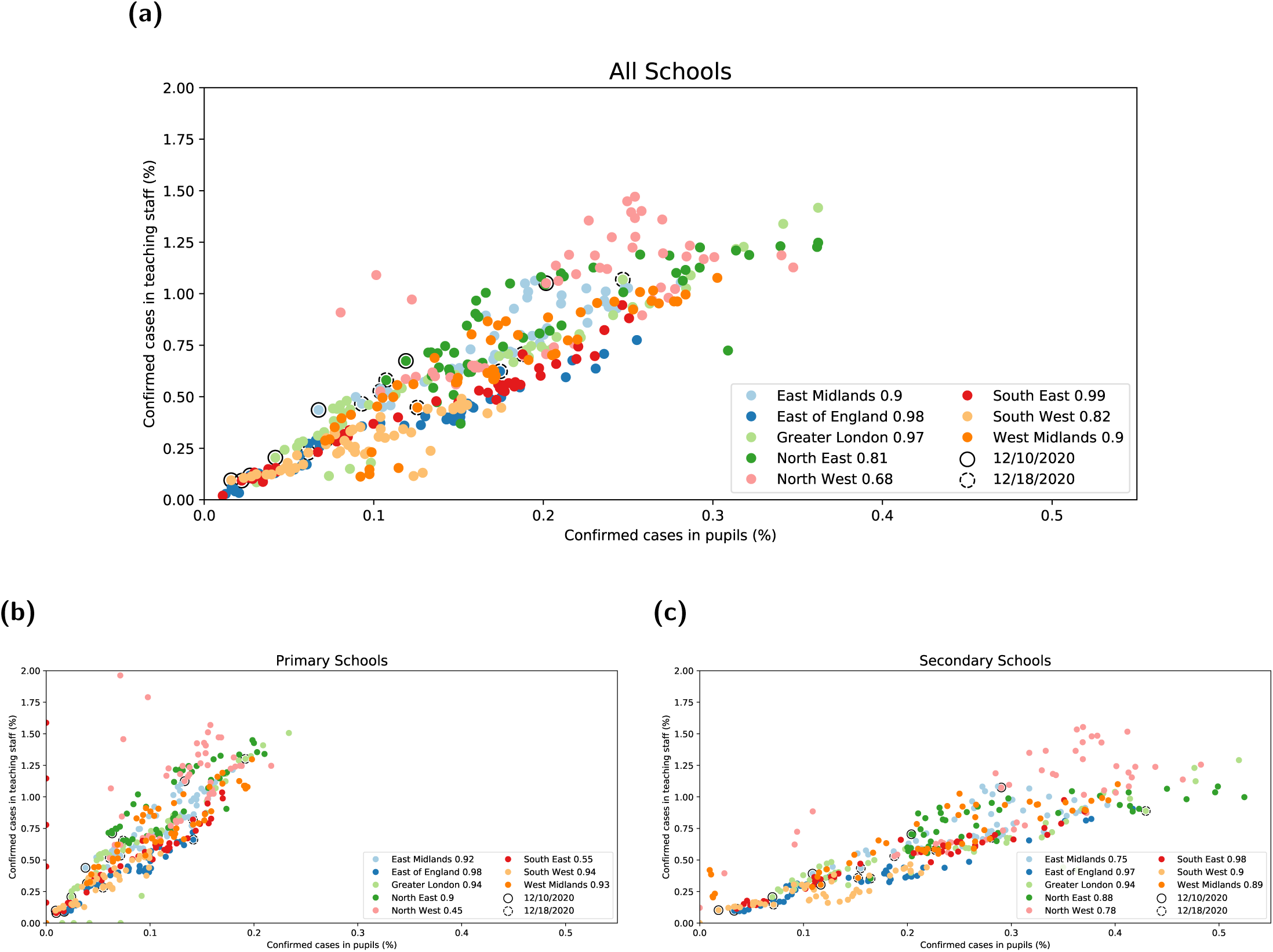
Confirmed cases in teaching staff (by percentage per region) against confirmed cases in pupils (by percentage per region) by day for all regions. For all panels, the solid circle for each region indicates the earliest date in this data set (14th October 2020) whilst the dashed circle indicates the latest date (18th December 2020). The correlation coefficient for each region is given in the legend. Cases are shown for all: **(a)** All schools; **(b)** primary schools only; **(c)** secondary schools only.

### 3.2 Analysis of absences owing to cases of COVID-19 in school children and teachers by tier status of LTLA of school location

We also examined the number of absences as a result of confirmed cases in pupils and teachers, stratified by tier status of the relevant local authority (Fig. 4). We observe a marked difference between students and teachers when stratified by tier status. In primary schools, cases in students increased slightly in tiers 1 and 2 for the first two weeks of the national lockdown in November, though remained relatively static in tier 3 (Fig. 4(a)). Cases then began to marginally reduce across all tiers. In secondary schools, confirmed cases in students increased across all tiers for the first two weeks of lockdown before decreasing (Fig. 4(b)). In tier 3 regions, cases continued to decline after lockdown, whilst there was a marginal increase in cases in tier 2 regions. We observe a different pattern of behaviour in teachers - confirmed cases in regions previously in tier 3 declined throughout the lockdown in both primary and secondary schools, whilst there was a marginal increase in confirmed cases in tier 2 and tier 1 regions during this same period (Fig. 4(c) and Fig. 4(d)). Cases in teachers increased slightly in tier 2 regions in the second week of December in both settings, whilst they continued to decline in tier 3 regions.

**Fig. 4:**
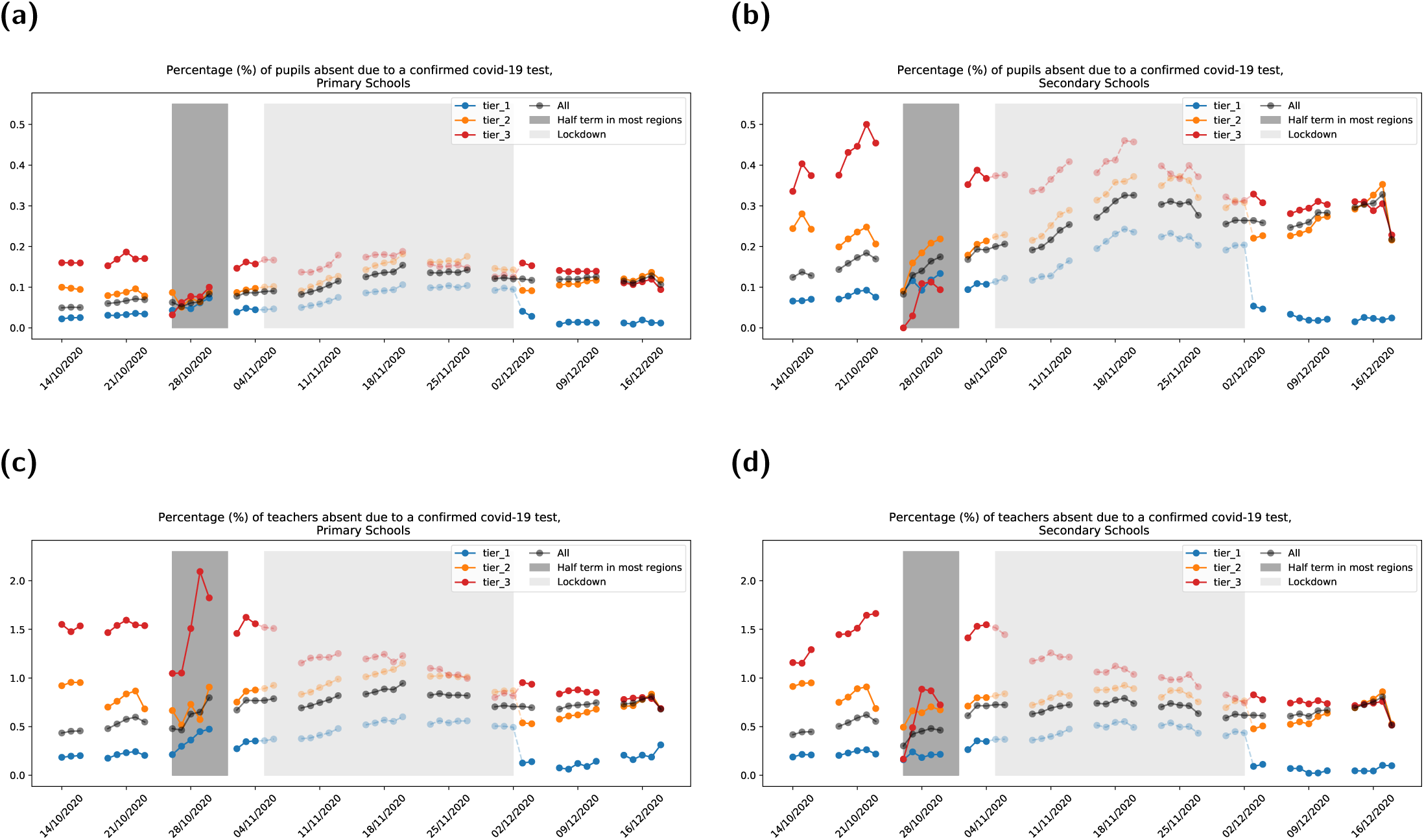
Percentage of study population recorded as a confirmed case, stratified by intervention tier status. For each panel, we display the number of cases by date and by intervention tier status, from 14th October 2020 to 17th December 2020. The half term week for most of England is shown by the dark grey shaded region, with the period corresponding to the national lockdown shown by the light grey shaded region. The faded dots indicate the tier status prior to the national lockdown that was introduced on Thursday 5th November 2020. **(a)** Pupils in primary schools. **(b)** Pupils in secondary schools. **(c)** Teachers in primary schools. **(d)** Teachers in secondary schools. It should be noted that several regions changed tiers when lockdown was lifted on 2nd December, leading to an observed discontinuity in the data displayed in the figure on this date.

### 3.3 Spatiotemporal analysis of community cases and cases in schools in November and December

We investigated the spatiotemporal behaviour of cases at the lower tier local authority (LTLA) level in the community, in school teachers and school pupils from early November to the end of term.

Community cases were highest in the North, the Midlands and Greater London at the start of the November lockdown. Cases were observed to decrease in these regions during lockdown with, in the last week of November, the emergence of a new cluster in the South East and London (Fig. 5 and Fig. 6, left columns). Cases in teachers (Fig. 5 and Fig. 6, middle columns) and pupils (Fig. 5 and Fig. 6, right columns) did reduce during lockdown, but at a slower rate. A new cluster of cases emerged in school teachers and pupils in late November, whilst the country was under lockdown and a similar increase in cases in the community is observed in December. At LTLA level there is some slight variation observed between local authorities reporting very high numbers of community cases and very high numbers of cases in schools.tea

**Fig. 5:**
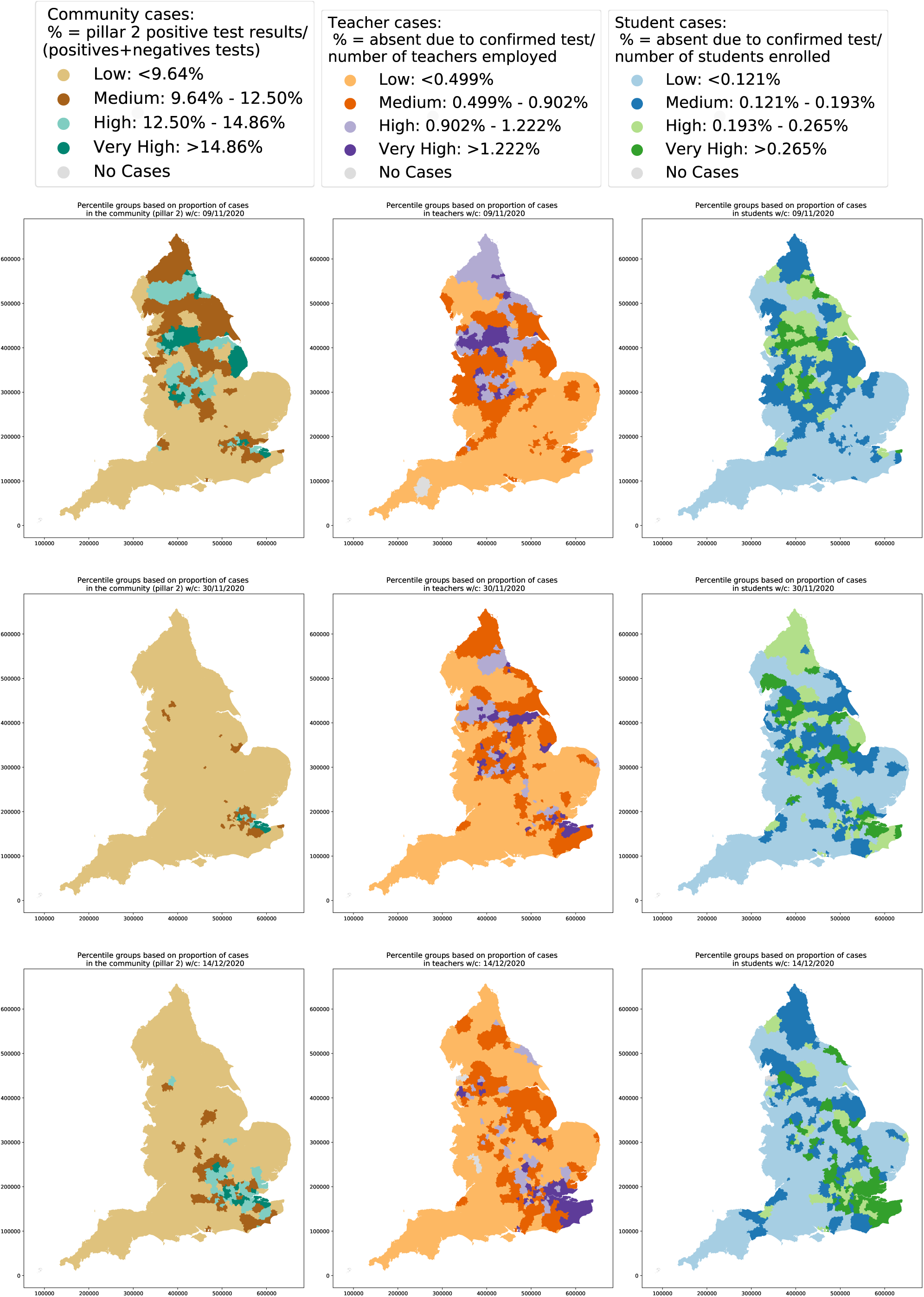
Relative incidence at LTLA level by week for England. For the week commencing 10th November, the top 10% of LTLAs are designated ‘Very high’, the 75th-90th percentile ‘High’, the 50th-75th percentile ‘Medium’ and the remainder ‘Low’. Cases are grouped into: **(left column)** community; **(middle column)** school teachers; **(right column**) school pupils.

**Fig. 6:**
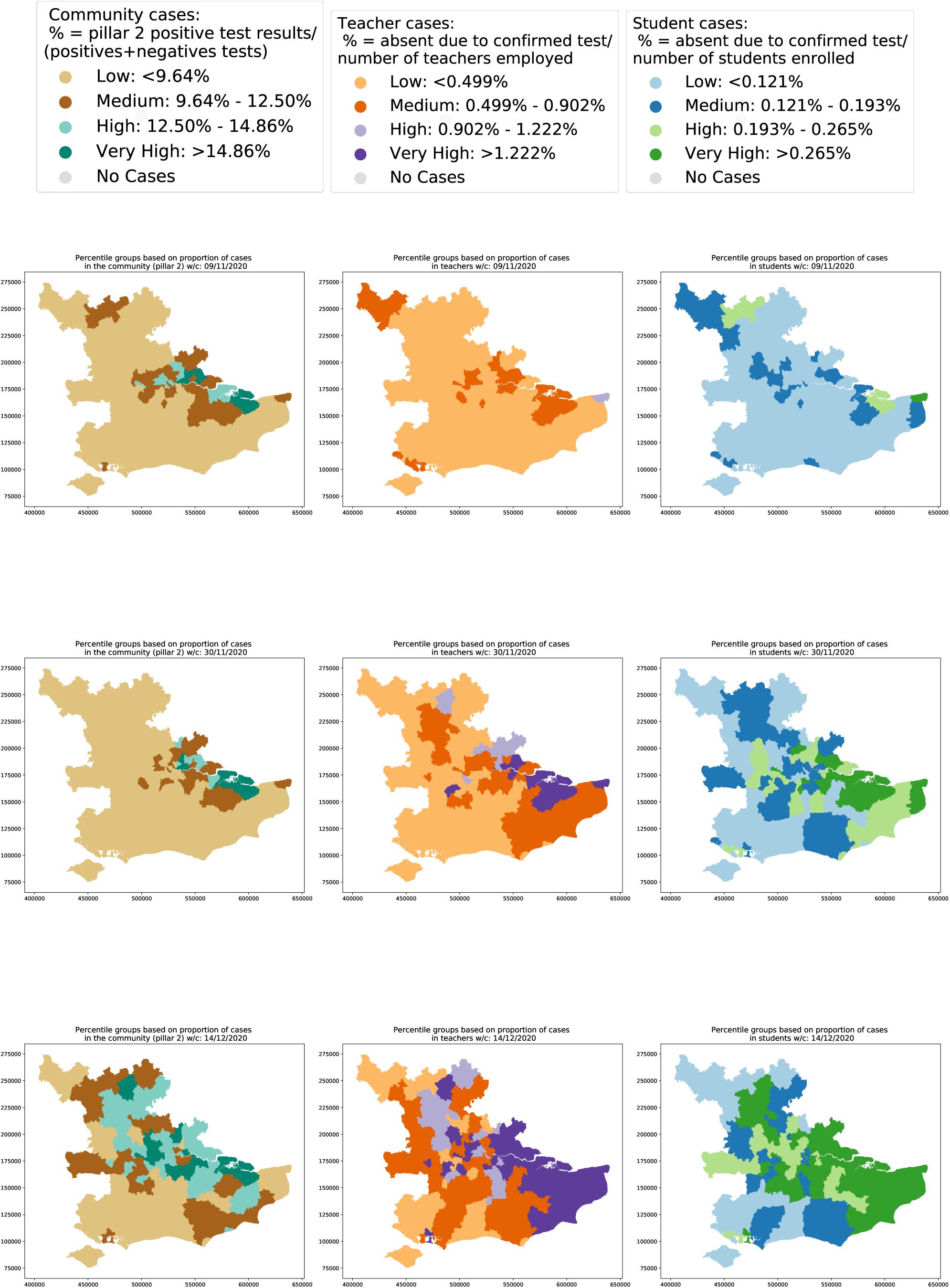
Relative incidence at LTLA level by week for the South East of England and Greater London. Thresholds to designate ‘Very high’, ‘High’, ‘Medium’ and ‘Low’ classifications are as defined in Fig. 5. Cases are grouped into: **(left column)** community; **(middle column)** school teachers; **(right column**) school pupils.

Given this observed increase in community cases in the South East during November and December, we investigated whether there was any signal indicating an increase in clusters of cases in schools during this period. We studied the frequency distribution of the number of confirmed cases by school on 4th November, the day before lockdown was introduced, and on 16th December, two days before the end of term (Fig. 7). We observe that, in Greater London and Kent, more secondary schools reported a greater number of students absent with confirmed infection in the last week of term compared to early November. However, we do not observe the same effect in primary schools in these regions - when absences with infection are reported in primary schools, in the majority of cases there is a single child absent with confirmed infection (Fig. 7, first row). When we compare this result with other regions such as Devon (Fig. 7, bottom left panel) and the West Midlands (Fig. 7, bottom right panel), we note that secondary school absences in the last week of term follow a similar distribution to early November.

**Fig. 7:**
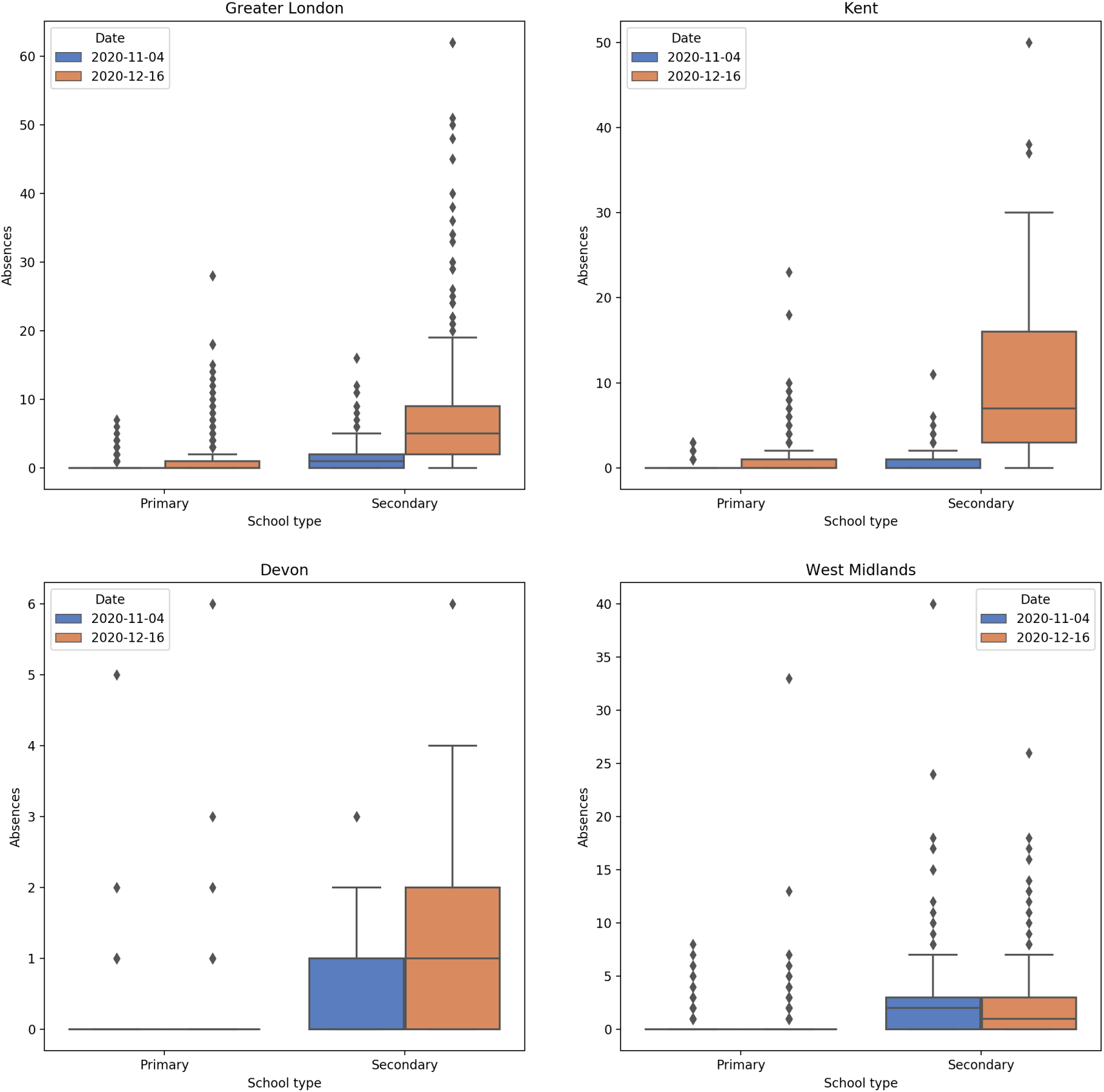
Frequency distribution of the number of absences due to a confirmed case per school. For all panels, box plots represent the distribution of absences due to a confirmed case in four regions of England. The four regions presented are: **(top left)** Greater London; **(top right)** Kent; **(bottom left)** Devon; **(bottom right)** West Midlands. On each subplot, we display the distributions for 4th November 2020 (blue) and 16th December 2020 (orange) for primary schools (first two plots) and secondary schools (second two plots).

Finally, we examined the temporal correlation at the LTLA level between cases in the community and cases in primary and secondary schools (Fig. 8). We varied the lag time between school and community cases to explore whether there was any signal that increased cases in schools resulted in increased cases in the community at a later time, or whether the opposite was the case. In London, Kent and the West Midlands, we observe a weak correlation between cases in secondary school pupils and community cases that increases with lag time, indicating that an increase in community cases is most positively correlated with an increase in school cases in pupils at a later date. We observe the same result for primary school pupils in Kent and the West Midlands, but noticeably observe a negligible correlation between community cases and cases in primary school children in London across all time lags. There is no clear signal in Devon, possibly owing to the relatively low number of cases observed in children in the county.

**Fig. 8:**
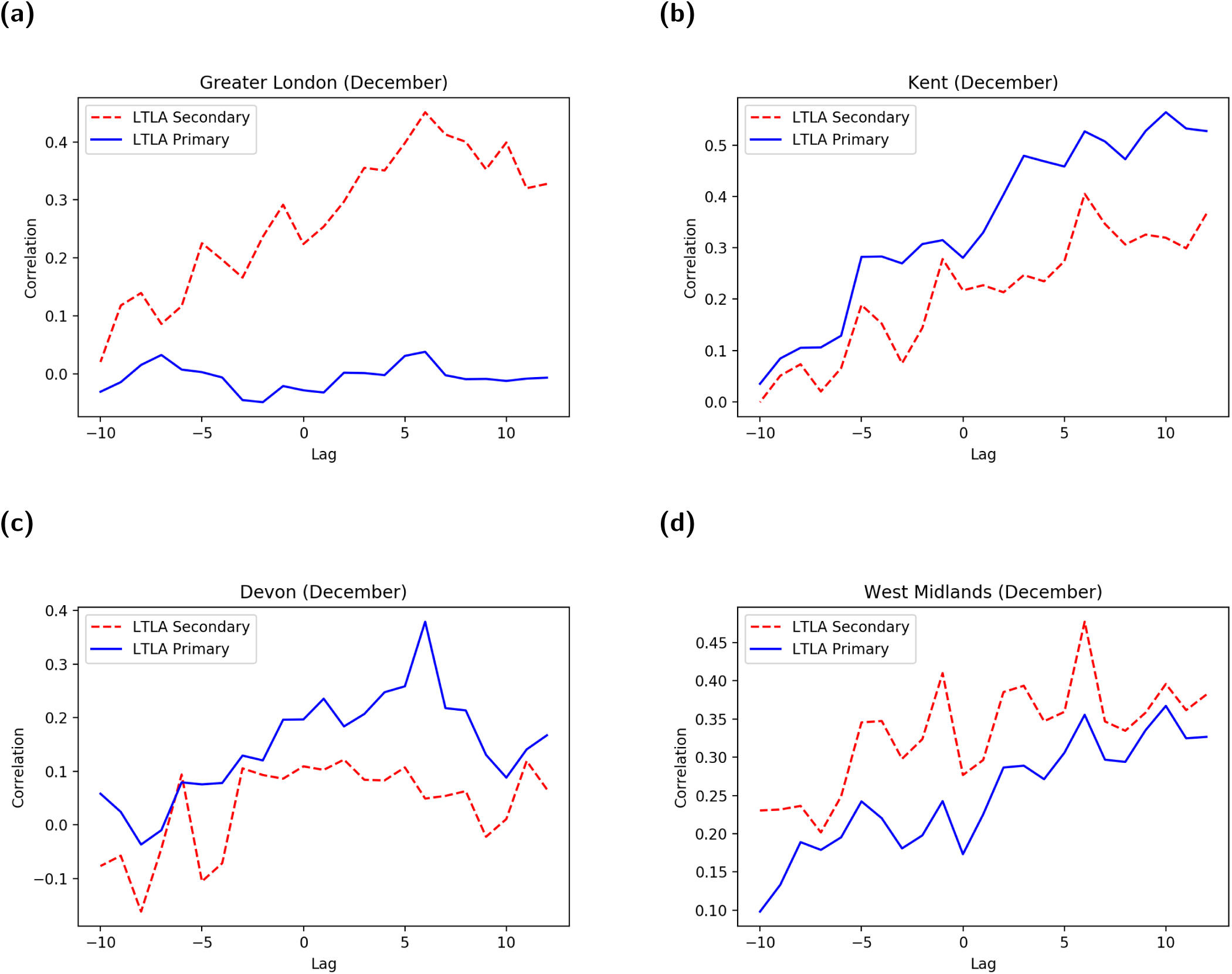
Correlation between cases in the community and pupils in December. In these panels, a positive lag indicates that the correlation is calculated between schools on the current date and community cases that have been reported at an earlier date up to a maximum lag of 12 days. The correlation is calculated for all LTLAs in each region, rather than calculated individually for each LTLA. For varying time lag applied to data from December, the LTLAs presented are: **(a)** Greater London; **(b)** Kent; **(c)** Devon; **(d)** West Midlands. Secondary schools are depicted by dashed red lines and primary schools by solid blue lines.

## 4 Discussion

In this paper, we present a set of analyses of the Department for Education data on Educational Settings recording school attendance, in order to investigate the impact of the pandemic upon schools and the potential role of school children upon transmission in the wider community. We observe that cases in schools increased throughout September and October 2020, mirroring the increases reported in the local community. The percentage of students with confirmed infection in secondary school students was found to be higher than in primary school students throughout this period. Notably, this was not the case with teachers - the percentage of teachers reporting infection appeared to be of a similar magnitude in both primary and secondary schools. This suggests that teachers are not exposed to increased risk in school environments where more children are infected, perhaps suggesting that the background incidence in the community plays a greater role in determining the risk to teachers. We can also infer that teachers are not at greater risk in primary schools than in secondary schools.

During the November 2020 lockdown, schools remained open and the observed rise in cases in younger people led to suggestions that schools were playing a major role in spreading the virus. However, the subsequent confirmation of the emergence of the more transmissible B.1.1.7 variant provided evidence to suggest that this may not be the case; the increase in cases in secondary school aged children in London, the South East and the East of England throughout late November and early December was not observed in the North West, the North East and the Midlands, where this new variant was not widely circulating at that point in time.

We seek to understand whether cases in schools are driving an increase in cases in the community, or whether an increase in incidence in the local area leads to increased infection rates in school-aged children and hence more cases reported in schools. Some insights can be gained by examining cases in schools stratified by the tier status of the relevant local authority. Notably, the increase in cases in students observed across all tiers during the first two weeks of the November lockdown, particularly in secondary schools, was not reflected in a rise in cases in teachers during this same period. If schools were exposing teachers to increased risk during this lockdown, we might expect that, as cases started to rise amongst secondary school children, a similar rise may be observed, following a time lag, in cases in teachers. Given that this is not the case, this may suggest that teachers are more at risk of infection in the community than in the school environment and the decreased community mixing due to the national lockdown led to the drop in cases in teachers during this period.

During December, we observed a distinct increase in the number of confirmed cases in students and teachers in the South East of England. However, this increase mirrored that seen in the local community. As the new variant B.1.1.7 became more prevalent, community cases increased more rapidly in the South East. We did observe some spatial variability at the LTLA level between areas of high incidence in the community and in schools. From our analysis, it appears that during December there was an increase in clusters of cases in secondary schools in those parts of the country that were most affected by the new variant. Kent in particular reported more schools with large numbers of students absent with confirmed infection in mid-December compared with before the start of the November lockdown. Noticeably, we did not observe a marked increase the number of students absent per school in primary schools in Kent. There has been much debate around the relative role of primary and secondary schools during the pandemic and this analysis at least suggests that primary school children do not appear to be as affected as secondary school children by the emergence of the new variant.

When we examined the relationship between community and school cases in more depth, we observed a correlation between cases in the community and cases in schools in most regions, with the strongest correlation between current cases in schools and community cases reported several days previously. From this analysis we conclude that there is not sufficient evidence to suggest that outbreaks in schools are driving an increase in community cases, with the calculated correlations providing weak evidence that suggesting the opposite may be true, that an increase in incidence in the community leads to more cases in schools. As schools re-open, careful monitoring may be required in order to determine the risk associated with open schools upon community incidence.

It is important to note that all of the data analysed here refer to absences in schools as a result of confirmed cases of COVID-19 in pupils and teachers, but they do not necessarily imply that these individuals were infected *within* schools. The data do not record location of infection and therefore we cannot provide conclusive evidence of the presence or absence of spread within a school.

At the time of writing at the end of January 2021, there have been almost 3.8 million confirmed cases of COVID-19 in the UK and around 105,000 deaths ([28]). Hospital occupancy is reaching capacity in many parts of the country, daily deaths are still above 1,000 per day and schools remain closed except for children of key workers and vulnerable children. It is clear that the longer that children remain out of school the greater the risk of many children suffering long term from a lack of access to face-to-face teaching and socialisation, with a resulting negative impact upon their mental health and education. It is vital that processes are put in place to ensure that children get back to school as rapidly but as safely as possible. Our work suggests that this can be achieved by ensuring that community incidence is as low as possible when schools re-open. However, further measures, such as ensuring parents do not mix at pick up and drop off and a reinforcement of the need for people to work from home if they can, may be needed in order for children to return to school safely in the near future.

## Data Availability

Data from the Department for Education Educational Settings database were supplied after anonymisation under strict data protection protocols agreed between the University of Warwick and the Department for Education in the UK. The ethics of the use of these data for these purposes was agreed by the Department for Education with the Government SPI-M(O)/SAGE committees.

## Author contributions

**Conceptualisation:** Alex Holmes; Emma Southall; Michael J. Tildesley.

**Data curation:** Alex Holmes; Emma Southall; Michael J. Tildesley.

**Formal analysis:** Alex Holmes; Emma Southall.

**Investigation:** Alex Holmes; Emma Southall.

**Methodology:** Alex Holmes; Emma Southall; Michael J. Tildesley; Matt J. Keeling; Edward Hill; Louise Dyson.

**Software:** Alex Holmes; Emma Southall.

**Validation:** Michael J. Tildesley; Matt J. Keeling; Edward M. Hill; Benjamin D. Atkins; Louise Dyson; Trystan Leng; Robin N. Thompson.

**Visualisation:** Alex Holmes; Emma Southall.

**Writing - original draft:** Michael J. Tildesley; Alex Holmes; Emma Southall, Edward M. Hill; Louise Dyson; Benjamin D. Atkins; Matt J. Keeling; Trystan Leng; Robin N. Thompson.

**Writing - review & editing:** Matt J. Keeling; Edward M. Hill; Louise Dyson; Benjamin D. Atkins; Trystan Leng; Robin N. Thompson; Alexander Holmes; Emma Southall; Michael J. Tildesley.

## Ethical considerations

The data were supplied by the Department for Education under strict data protection protocols agreed between the University of Warwick and the Department for Education. The ethics of the use of these data for these purposes was agreed by the Department for Education with the Government’s SPI-M(O) / SAGE committees.

## Data availability

Data from the Department for Education Educational Settings database were supplied after anonymisation under strict data protection protocols agreed between the University of Warwick and the Department for Education in the UK. The ethics of the use of these data for these purposes was agreed by the Department for Education with the Government’s SPI-M(O)/SAGE committees.

